# Nutritional status and health of community-dwelling elderly people in urban areas of the Lalitpur district, Nepal

**DOI:** 10.1101/2025.10.03.25337277

**Authors:** Eebaraj Simkhada, Aerusha Simkhada

**Affiliations:** Eebaraj Simkhada, Department of Internal Medicine, Manmohan Memorial Teaching Hospital, Nepal; Aerusha Simkhada, Aditya College of Pharmacy, Jawaharlal Nehru Technological University, Andhra Pradesh, India

**Keywords:** MNA, Nepal, Elderly, Urban, Nutritional Assessment, BMI

## Abstract

**Background:** Poor nutrition causes ill health and functional dependence in the older population. Different aspects of elderly health and nutritional status are little known in the growing elderly population in Nepal. This study describes the nutritional status and its association with socio-demographic factors, and health-related characteristics among community-dwelling elderly people in urban areas of Lalitpur district of Nepal.

**Methods:** The study was a cross-sectional population-based study among elderly people aged 60 years and older, in 5 VDCs around the outskirts of Lalitpur Metropolitan City of Lalitpur district of Nepal. Multi-stage cluster sampling with probability proportional to size was used to select a sample of 360 elderly individuals. Nutritional status was assessed by the Mini Nutritional Assessment (MNA) tool. The data on socio-demographic factors, socioeconomic factors, and health characteristics were presented in percentages, means, and standard deviations. Chi-square, and ANOVA tests were performed to assess the associations of variables with malnutrition.

**Results:** Using the MNA score, this study found 13% malnutrition and 45% at risk of malnutrition among participants (177 men and 183 women with a mean age of 71.2 years). About 28% of the study population was overweight with body mass index (BMI) > 25.0. About one-fourth (25.6%) were totally dependent financially on their children. Approximately 65.3% were diagnosed with chronic diseases. Hypertension was the most prevalent disease (26.5%), followed by Diabetes Mellitus. About 28.6% reported insomnia and 18.6% of individuals reported having chronic pain. Educational status, agriculture as an occupation, increased elderly age, and having insomnia were significantly associated with nutritional status.

**Conclusions:** About half of the population had poor nutritional status. Elderly people were suffering from chronic diseases, chronic pain, insomnia, and financial dependency. This study indicates an urgent need to take care of the health of the elderly with timely intervention at the community and national level.

## Introduction

Nutrition is crucial for the health and development of an individual (1). Poor nutritional status is a common problem in older people. The well-being of the elderly adult can be adversely affected by malnutrition, causing a steady decline in functional status and susceptibility to complications associated with chronic illnesses inherent in this age group. Malnutrition potentially impacts morbidity, mortality, and quality of life in the elderly (2).

An estimate indicated that about 805 million people were chronically undernourished in 2012–14 (3). Undernutrition is highly prevalent in developing regions and older adults are more likely to be affected by malnutrition than young persons.

Poor nutrition causes ill health and functional dependence in the older population. Increased susceptibility to complications associated with chronic illnesses is common in this age group. In older individuals, poor financial condition, having multiple comorbidity, reporting chronic pain, gender (women being at higher risk), and having mental disorders were implicated in malnutrition (5).

Poor nutritional status is associated with an increased risk of cardiac and respiratory problems, infections, deep vein thrombosis and pressure ulcers, peri-operative mortality, and multi-organ failure (6). It has been implicated in the development and progression of chronic diseases commonly affecting the elderly, including osteoporosis, diabetes mellitus, and cancer (6,7).

Thus, malnutrition is considered the greatest threat to the health, well-being, and autonomy of elderly people (8).

The population of the world is becoming older as a consequence of a decline in fertility and increased life expectancy. It is projected that by the year 2050, the population of persons over 60 years will be two billion (17). Aging should be taken as a privilege and a societal achievement. However, national health policies and programs give less priority to the health of the elderly population.

Developing countries are aging faster than developed countries. As in most other countries, the proportion of elderly people is increasing in Nepal with annual elderly population growth rate of 3.39% (4). Indeed, population aging can have major implications on health and social services, as increased age results in a greater likelihood of having a disability and needing assistance (23), since malnutrition is a risk factor for the health, wellbeing, and autonomy of old people.

In a study, elderly persons, health problems and increased age had a negative impact on nutritional status. Educational status and expenditure on food were directly associated with poor nutritional status among the elderly (9).

A study in Lebanon (10) concluded that poor nutritional status was common in elderly women as compared to males. Women were socioeconomically disadvantaged, and they reported lower self-reported health status. As compared to males, female elderly people had high comorbidity, high drug intake, higher frailty, and disability.

Edentulous elderly having problems in mastication was associated with poor nutritional status in a study (11). Chronic pain is also a significant factor for malnutrition. Pain, fatigue, illnesses, and gastrointestinal problems may also decrease appetite and cause poor nutrition (5).

The major concern for the elderly are increased longevity with the rise of comorbidity, decline in physical ability and loss of cognitive function (12). Aging is a process in which an individual’s physical and mental abilities are gradually lost, and vulnerability to diseases increases in old age. Dietary habits and low-nutritious diets are also implicated for comorbidity.

Living alone is a risk factor for malnutrition, as demonstrated in a previous study (18). In old age, more elderly people become financially dependent on their children. The financial burden is also increased by co-morbidity, frequent hospitalization, and regular drug use.

Nutritional status of the elderly is influenced by a complex interaction of dietary, socioeconomic, physical, and psychological factors (21). Malnutrition will limit daily activities and worsen co-morbidities (12). The health of elderly people is largely affected by lifestyle, occupation, nutrition, chronic diseases, environmental factors, and psychosocial characteristics (22).

Increased co-morbidities are independently associated with poor nutritional status. Malnutrition in old age often goes undetected, and timely nutritional screening is crucial for early intervention.

Information about the nutritional status and health status of the elderly will be of public health importance because of the growing elderly population, and different aspects of elderly health, socioeconomic status, and social indicators need to be explored. A status report published by the Nepal Geriatric Centre in 2010 highlighted the research gap regarding health status and nutrition (23) and recommended research in the area in Nepal.

Reviewing 25 previous studies mostly from developed countries (n=14,149) on the nutritional status of the elderly population, Guigoz et al (24) reported a mean prevalence of malnutrition among community-dwelling elderly that ranged between 0 and 8%, and at risk of malnutrition of 45%. A higher prevalence of malnutrition was found in hospitalized elderly patients (20%), and elderly people residing in institutional homes (37%). Domit et al (25), in a study on nutritional and health status among nursing home residents in Lebanon, reported a gender difference in health, socioeconomic factors, and nutritional status. The malnutrition rate was 3.2%, and the at-risk-of-malnutrition rate was 27.6%. A study conducted in Nepal found that 31% of elderly people in Pharping VDC, near Kathmandu were malnourished, and 51% were at risk of malnutrition. This prevalence is higher in community-dwelling older adults than in studies conducted in India (26) and Bangladesh (9).

Although malnutrition is more prevalent in low-income countries where health conditions are worse among the elderly due to low socioeconomic status and health-related factors (10), unfortunately, little is known about the characteristics and the needs of these elderly people in Nepal. Situation of elderly physical health, mental health, and social health in Nepal is largely unexplored. Like many areas of public health, substantial constraints exist on the availability and quality of information to describe how big the problem of malnutrition is in the elderly and what the risk factors associated with the problem are in Nepal. Studies to explore the various aspects of health characteristics, social indicators, and nutritional status of the elderly population in Nepal are very limited.

Although research has shown that old age itself, gender, educational status, poverty, comorbidity, decreased cognition, depression, and social isolation are associated with poor nutritional status, the contribution of various factors to increasing the vulnerability of poor nutritional status among the elderly population in developing countries like Nepal may not be well recognized.

An increased understanding of the factors that contribute to poor nutrition in the elderly should enable the development of appropriate preventive and treatment strategies and improve the health of older people. This study is aimed at investigating the socioeconomic factors, physical health characteristics, and nutritional status among community-dwelling elderly people in urban areas of the Lalitpur district, Nepal.

## Methods

### Study design

The design of the study was a cross-sectional population-based study among elderly people aged 60 years or older in urban areas of Lalitpur district. A sample of 360 elderly individuals from areas located on the outskirts of the Lalitpur Metropolitan City was taken for the study.

### Sample size

The sampling technique used was multi-stage cluster sampling with probability proportional to size. The sampling technique used was multi-stage cluster sampling with a probability proportional to size. The sample size was established according to the prevalence of malnutrition among home-living elderly people. A review of the literature on MNA for the prevalence of malnutrition among the community-dwelling elderly population (21 studies) revealed 2% malnutrition and 24% at risk of malnutrition in developed countries (24). Studies among hospital outpatients and home care elderly showed a higher prevalence of malnutrition (9%) and at risk of malnutrition (45%) (24). A study found 26% malnutrition among rural elderly people in Bangladesh using MNA to assess nutritional status (9). In a VDC near Kathmandu Valley, using MNA to assess malnutrition found that 31% of the elderly population were malnourished and 51% were at risk of malnutrition (36).

The sample size was calculated through Epi Info version 7 (Statcalc) and using the formula of a population survey with the following values:

Taking an expected frequency of 31% with a confidence limit of 7% and a design effect of 2, from 30 clusters, 360 participants were necessary to establish a 95% confidence interval.

### Study Setting

The study site was the surrounding areas of Lalitpur Metropolitan City of Lalitpur District, Nepal. This study used the old administrative division of Lalitpur District prior to the federal restructuring of Nepal for the sampling purpose and mentioned former VDCs were later merged as municipalities in 2017 (16).

The district is divided into three electoral constituencies. Densely populated Electoral Constituencies No. 2 and 3, consisting of 13 VDCs, were selected purposively. From the 13 VDCs, 5 VDCs (Lele, Siddhipur, Thaiba, Saibu, and Khokana) were selected randomly. Each VDC was divided into 9 wards, and each ward was taken as a cluster. In total, 45 clusters from 5 VDCs, 30 clusters were selected according to probability proportionate to size. Within each cluster, 12 elderly individuals were selected randomly.

**Fig. 1:**
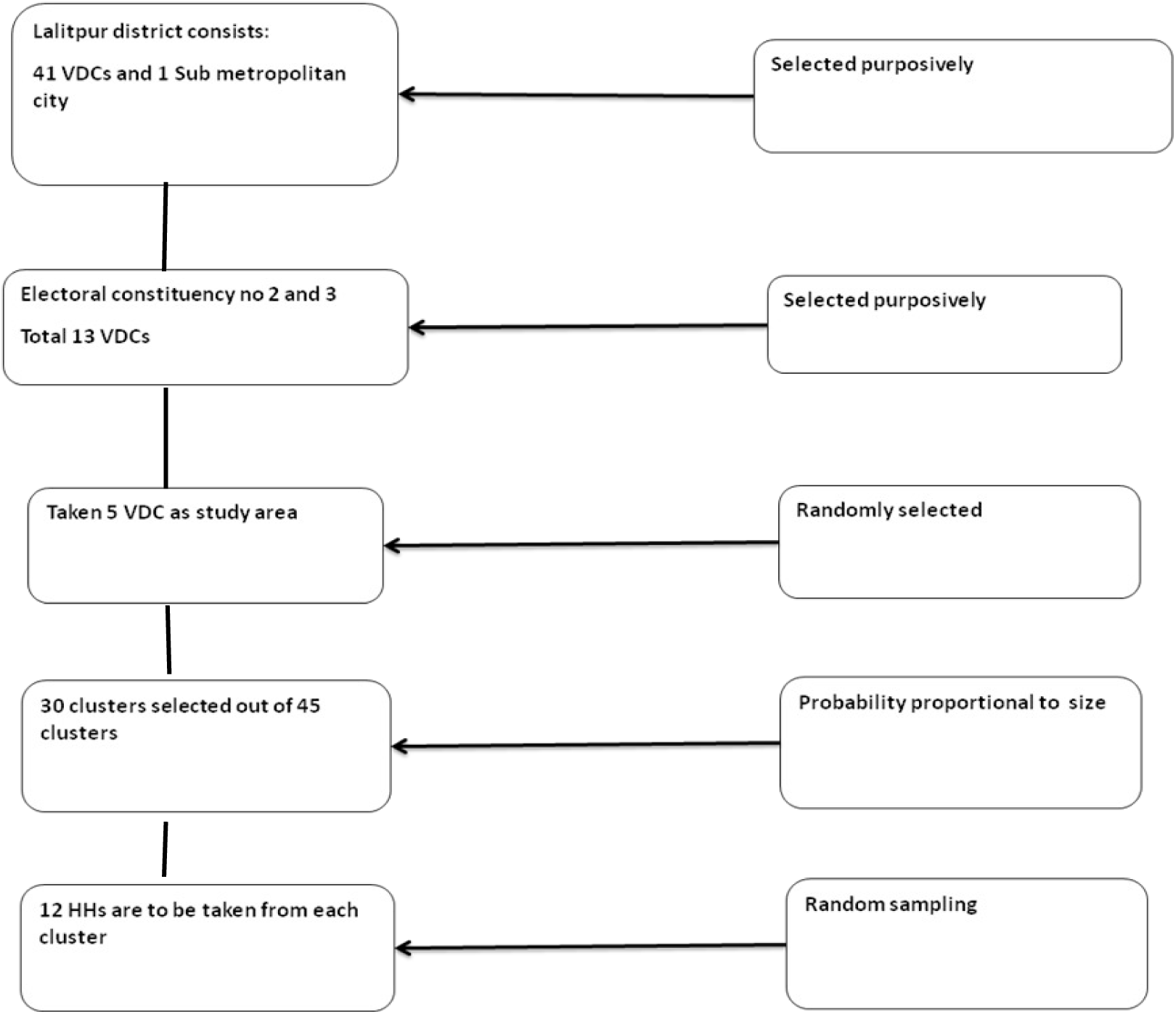
Sample selection flow chart

Only one elderly person was selected from each household based on their willingness to participate and availability. In case of refusal to participate or unavailability at the time of data collection, another elderly person from an adjacent household was selected as a replacement. The inclusion criteria of the study were participants aged 60 years or older who consented to participate in the study and did not have a severe mental illness.

### Ethics and consent

Ethical approval was obtained from the Institutional Review Committee (IRC) at the Institute of Medicine, Maharajgunj Medical Campus, in Kathmandu, Nepal. The purpose of the study and data collection, including anthropometric measurements, were explained to the participants. Written consent was obtained from all participants. Participants were fully informed of their rights to decline or withdraw from participation in the study if desired. The information collected was kept confidential.

### Study tools

#### Study questionnaire

A multi-component structured questionnaire including assessment tools was used to collect data in the study. The interviewers consisted of three trained personnel, having previous experience in field surveys and research. The questionnaire was translated into the Nepali language from English and then back-translated into English by two persons fluent in both languages. Pre-testing of the questionnaire was done in a sample of 30 elderly people to identify the feasibility of the questionnaire. According to the feedback, the questionnaire was revised, and minor changes were made. Written consent was taken prior to the interview at their place of residence from all participants. Each interview took 35 minutes on average. In case of the inability to communicate effectively due to language or speech leading to difficulty in understanding, the help of a family member was taken. The survey was conducted over 48 days, from September 20th to November 7th, 2014.

#### Assessment tools

##### Sociodemographic factors

Demographic characteristics included in the study were age, gender, ethnicity, religion, place of residence, marital status, and living condition (living alone or living with others). Socioeconomic status was assessed by a weighted wealth index method used by the Department of Health Survey (28). The index assessed housing characteristics including floor, wall, and roof materials; ownership of agricultural lands; and household assets including television, electricity, telephone, motorcycle, fan, refrigerator, and computer in the house. Availability and use of drinking water for the household, use of fuel for cooking, and use of a toilet were also assessed by the index. All the items were dichotomized, and factor analysis was done by principal component analysis to reduce the items and extract factor 1. This procedure first standardized the indicator variables; then, the factor coefficient scores (factor loadings) were calculated. For each household, the indicator values were multiplied by the factor loadings as item weights and summed up to produce the household’s wealth index value. Then, the total wealth index score was divided into five categories: lowest, second, middle, fourth, and highest income groups.

Ethnicity was classified into groups: Brahmin/Chhetri, Newar, other Janajati (Magar, Tamang, Gurung), and Dalit.

Financial dependency on children was assessed by asking a question with three answers: “totally dependent,” “partially dependent,” and “independent.” Education was categorized as: 1) illiterates (unable to read and write), 2) informally literate, 3) Primary education, and 4) Secondary education and above (included those who had completed primary education, those having SLC, and a university degree). Main occupation was recorded based on current occupation as farmer, laborer, self-employed/job, and housework/no work. Type of family was categorized into 3 groups: nuclear, joint, and extended family.

##### Nutritional status and anthropometric measures

To assess nutritional status, the Mini Nutritional Assessment (MNA) was used. It is an instrument that allows for an objective assessment of nutritional status in older people. This assessment tool is widely used in elderly people and has been translated into more than 20 languages. It is a well-validated tool cited in various publications.

The MNA includes 18 questions grouped into 4 parts:

1) Anthropometric assessment: BMI calculated from weight and height, weight loss, and arm and calf circumferences; 2) General assessment: lifestyle, medication, mobility, and presence of signs of depression or dementia; 3) Short dietary assessment: number of meals, food and fluid intake, and autonomy of feeding; 4) Subjective assessment: self-perception of health and nutrition. Each answer has a score, and the maximum MNA score is 30. The MNA was first translated into Nepali carefully to preserve the original meaning of the MNA questions and revised by the researcher and field interviewers. The MNA is a nutritional status assessment tool with a reliable scale and clearly defined thresholds. The MNA score of each participant was calculated, and scores were categorized as < 17 points for malnutrition, 17-23.5 for nutritional risk, and ≥ 24 for well-nourished.

Body weight was measured in light clothes and no shoes by electronic digital scale from Microlife company-Switzerland (model no. WS 50) to the nearest 0.1 kg, and height was measured with a measuring tape mounted on a wall to the nearest 0.5 cm. Measurement was taken in standing position without shoes, feet put together with heels, buttocks, shoulder, and back of the head touching the wall, and head facing straight forward. Body mass index (BMI) was calculated with the formula: Weight (kg) / height (m^2^). Mid-upper arm circumference was measured with a non-elastic tape on the relaxed arm. Calf circumference was measured with a non-elastic tape on the thickest part of the undressed calf, with the individual sitting and their knee flexed at a 90-degree angle.

##### Health characteristics

Health status was assessed by self-reported health (SRH). SRH was measured by asking the question, “In general, how do you describe your health?” Answers ranged between poor health, fair health, and good health. Self-reported co-morbidities were recorded based on physician-diagnosed chronic diseases (hypertension, diabetes, heart disease, stroke, cancer, arthritis, liver or gall bladder problem, respiratory problem, and no chronic diseases). Responses were again recorded in categories: No chronic illness, one chronic illness, two chronic illnesses, and three or more chronic illnesses. Daily drug intake was recorded as the number of drugs taken daily as prescribed by a physician. Chronic pain, defined as participants feeling pain for at least 3 months, was determined by asking a yes/no question. Participants were asked about insomnia and responses were categorized into two: no or occasionally, and often or always. Recent hospitalization, defined as hospitalization within 1 year, was recorded as yes or no.

Oral health status was assessed by yes/no questions in three domains: having chewing problems, having loss of dentition (no loss, partial loss, and total loss), and wearing dental prostheses. Tobacco smoking was assessed by a yes/no question asking whether the elderly participant was currently smoking or not smoking. Current smoking was defined as the participant having smoked within the past month. Physical exercise was assessed by asking if the individual had been exercising daily for 30 minutes for over a year.

### Statistical analysis

Data editing and coding was done by the researcher every day. Data entry was done in EpiData 3.1.

Data analyses were carried out using the Statistical Package for the Social Sciences (SPSS) software, version 20.0. Percentages were used to present nominal variables, while means and standard deviations were calculated for continuous variables. Chi-square tests were performed to assess associations between independent variables (socio-demographic factors, health characteristics) and nutritional status. An ANOVA test was performed to compare means.

Validated instruments were used. Probing questions were asked. MNA was used in previous studies in Nepal and showed good reliability (13, 36). Reliability was assured by translation/re-translation of tools (English-Nepali-English). Study tools and instruments were pre-tested at Tikathali VDC, Lalitpur, Nepal. Training of the enumerators for two days was done.

## Results

From multistage cluster sampling, a sample of 360 participants was selected for the study. Among the selected elderly people, 6.11% refused to participate in the study and were replaced. The age and gender of the participants who refused and those who were replaced did not differ significantly.

### Sociodemographic characteristics

Table 1 presents the sociodemographic characteristics of the elderly people. The study sample included 183 women (50.8%) and 177 men (49.2%). The mean age was 71.2 (SD 8.2) years and was similar in both genders. Nearly 90% of the respondents were Hindu by religion, 7.2% were Buddhists, and 1.9% were Christians. Ethnicity wise, nearly 68% of the participants were Newar, 26.9% were Brahmin/Chhetri, 3.9% were other Janajatis including Tamang, Magar, and Gurung, and 1.7% were Dalits.

**Table 1.** Socio-demographic characteristics of the study sample.

| Variables |  | Frequency<br>(n=360)<br>n (%) | 95% CI |
| --- | --- | --- | --- |
| Age class | 60-70 | 200 (55.6) | 49.5-60.8 |
|  | 71-80 | 101 (28.1) | 23.3-33.1 |
|  | >80 | 59 (16.4) | 12.8-20.0 |
| **Gender** | Male | 177 (49.2) | 44.4- 53.9 |
|  | Female | 183 (50.8) | 46.1- 55.6 |
| **Ethnicity** | Newar | 243 (67.5) | 63.1-72.5 |
|  | Brhamin/Chhetri | 97 (26.9) | 21.7-31.4 |
|  | Janjati (other) | 14 (3.9) | 1.9-5.8 |
|  | Dalit | 6 (1.7) | 0.6-3.1 |
| **Marital status** | Married | 212 (58.9) | 53.9-63.9 |
|  | Widowed | 148 (41.1) | 36.1-46.1 |
| **Education** | No education | 244 (67.8) | 63.6-72.2 |
|  | Literate(informally) | 52 (14.4) | 10.8-17.8 |
|  | Primary school | 25 (6.9) | 4.7-9.7 |
|  | Secondary and above | 39 (10.8) | 8.1-14.2 |
| **Main occupation** | Agriculture | 258 (71.7) | 67.5-76.9 |
|  | Employed/business | 51 (14.2) | 10.6-18.1 |
|  | Housework | 51 (14.2) | 10.8-17.5 |
| **Living condition** | Living alone | 3 (0.8) | 0.0-1.7 |
|  | Living with other | 357 (99.2) | 98.3-100.0 |
| **Financially dependent on children** | No | 131 (36.4) | 30.8-40.5 |
|  | Partially | 137 (38.1) | 33.3-42.8 |
|  | Totally | 92 (25.6) | 21.4-30.6 |

Nearly 59% were currently married, and 41.1% were widowed. Among the participants, 67.8% were illiterate, 14.4% did not have formal education but were literate, 6.9% had completed primary level education, and 10.8% had secondary level or higher education. About 85.8% of elderly people were living in a joint family, and 0.8% of the participants were living alone. Regarding main occupation, 71.7% were farmers by occupation, 14.2% were employed or having their own business, and another 14.2% had no job, doing housework.

### Nutritional status and health related characteristics

Table 2 shows the nutritional status and health-related characteristics of the study population. According to MNA, 13.1% of the study population were malnourished (MNA score < 17). The mean MNA score was 22.15 (SD=4.23). The mean BMI was 23.09 (SD=4.07). The underweight elderly population had a BMI< 18.5.

**Table 2.** Nutritional status and health related characteristics of the study sample.

| Variables |  | Total (n=360)<br>n (%) | 95% CI |
| --- | --- | --- | --- |
| **MNA** |  |  |  |
| **Mean(SD)** |  | **22.15 (4.23)** |  |
|  | Malnutrition (<17) | 47 (13.1) | 9.7-16.4 |
|  | At risk of malnutrition (17-23.5) | 162 (45.0) | 40.3-50.6 |
|  | Normal (>24) | 151 (41.9) | 36.7-47.2 |
| **BMI** |  |  |  |
| **BMI mean (SD)** |  | **23.09(4.07)** |  |
|  | Underweight (<18.5) | 42 (11.7) | 8.3-15.0 |
|  | Normal weight (18.5-24.99) | 216 (60.0) | 54.4-64.7 |
|  | Over weight (25-29.99) | 84 (23.3) | 19.2-28.1 |
|  | Obesity (>30) | 18 (5.0) | 3.1-7.2 |
| **Self rated health(SRH)** |  |  |  |
|  | Good | 121 (33.6) | 28.3-38.3 |
|  | Average | 208 (57.8) | 52.5-63.3 |
|  | Poor | 31 (8.6) | 6.1-11.7 |
| **Number of chronic diseases** | <=2 | 384 (96.7) | 94.7-98.3 |
|  | >2 | 12 (3.3) | 1.7-5.3 |
| **Daily drug intake** | <=3 | 316 (87.8) | 84.2-90.8 |
|  | >3 | 44 (12.2) | 9.2-15.8 |
| **Chronic pain** | Yes | 67 (18.6) | 14.7-22.8 |
|  | No | 293 (81.4) | 77.2-85.3 |
| **Hospitalization during last year** | Yes | 48 (13.3) | 9.7-16.7 |
|  | No | 312 (86.7) | 83.3-90.3 |
| **Insomnia** | Always/Often | 103 (28.6) | 23.9-33.6 |
|  | Occasionally/No | 257 (71.4) | 66.4-76.1 |
| **Regular Physical exercise** | Yes | 90 (25.0) | 20.6-28.6 |
|  | No | 270 (75.0) | 71.4-79.4 |
| **Current smoker** | Yes | 122 (33.9) | 28.9-38.9 |
|  | No | 238 (66.1) | 61.1-71.1 |

The 5-item self-rated health (SRH) scale was categorized into poor health (bad or very bad health), fair health (average health), and good health (very good or good health). One-third of the participants reported their health status as good health. Nearly 4% of the individuals reported having more than two chronic illnesses. About 37.8% reported having no chronic diseases, and 3.3% had more than two chronic diseases. Nearly 59% reported one or two chronic diseases. About 34.7% were not having any diagnosed chronic diseases. Hypertension was the most prevalent disease (26.5%), followed by Diabetes Mellitus (12.5%), COPD/Asthma (12.0%), arthritis (10.5%), CVD (2.3%), stroke (1.0%), and cancer (0.5%).

Nearly 34% were currently smoking tobacco. Regarding physical exercise, 25% of the participants were regularly exercising, and there was no significant difference between men and women on smoking and physical exercise.

About 12.2% of individuals reported taking more than 3 prescribed medicines daily. About 18.6% of individuals reported having chronic pain (feeling pain for at least 3 months). About 28.6% reported having insomnia always or often. About 13.1% of the individuals were hospitalized in 1 year.

Regarding oral health status, 46.4% reported having a problem chewing, and 83.6% were partially or totally edentulous.

### Association of nutritional status with socioeconomic and health characteristics

The purpose of this section is to examine the association between the independent variables and the dependent variable, i.e., nutritional status.

Table 3 shows the results of sociodemographic factors and their association with nutritional status. Age was significantly associated with poor nutritional status (p=0.04). No significant association was demonstrated between independent variables such as gender, ethnicity, living condition, marital status, and place of residence.

**Table 3.** Nutritional status and associated sociodemographic factors.

| **Variables** | **Total**<br>**(n=360)** | **Malnourish**<br>**ed**<br>**47, 13.1%** | **Risk of**<br>**malnutrition**<br>**162,45.0%** | **Well**<br>**nourished**<br>**151,41.9%** | **Test**<br>**statistics** | **p-**<br>**value<sup>a</sup>** |
| --- | --- | --- | --- | --- | --- | --- |
| **MNA mean (SD)** | 22.15(4.23) | 14.56(2.36) | 20.80(1.92) | 25.97 (1.62) | 747.41<sup>b</sup> | <0.001<sup>a</sup> |
| **BMI mean (SD)** | 23.09(4.07) | 18.92(2.52) | 22.40(3.98) | 25.13 (3.24) | 166.92<sup>b</sup> | <0.001<sup>a</sup> |
| **Age mean (SD)** | 71.21(8.25) | 73.00(9.21) | 71.60(8.59) | 70.21(7.45) | 2.43<sup>b</sup> | 0.088<sup>a</sup> |
|  |  | **n (%)** | **n (%)** | **n (%)** |  |  |
| **Age class, n(%)** |  |  |  |  |  |  |
| 60-70 | 200 | 19(9.5) | 88(44.0) | 93(46.5) | 9.94 | 0.041 |
| 71-80 | 101 | 16(15.8) | 43(42.6) | 42(41.6) |  |  |
| >80 | 59 | 12(20.3) | 31(52.5) | 16(27.1) |  |  |
| **Gender, n(%)** |  |  |  |  |  |  |
| Male | 177 | 26(14.7) | 72(40.7) | 79(44.6) | 2.75 | 0.252 |
| Female | 183 | 21(11.5) | 90(49.2) | 72(39.3) |  |  |
| **Ethnicity n(%)** |  |  |  |  |  |  |
| Brhamin/Chettri | 97 | 15(15.5) | 47(48.5) | 35(36.1) | 9.02 | 0.172 |
| Newar | 243 | 29(11.9) | 103(42.4) | 111(45.7) |  |  |
| Janjati (other) | 14 | 1(7.1) | 10(71.4) | 3(21.4) |  |  |
| Dalit | 6 | 2(33.3) | 2(33.3) | 2(33.3) |  |  |
| **Living status n(%)** |  |  |  |  |  |  |
| Living alone | 3 | 0(0) | 2(66.7) | 1(33.3) | 0.76 | 1.000 |
| Living with other | 357 | 47(13.2) | 160(44.8) | 150(42.0) |  |  |
| **Marital status n(%)** |  |  |  |  |  |  |
| Married | 212 | 26(12.3) | 90(42.5) | 96(45.3) | 2.36 | 0.307 |
| Widowed | 148 | 21(14.2) | 72(48.6) | 55(37.2) |  |  |
| **Education n(%)** |  |  |  |  |  |  |
| No education | 224 | 31(12.7) | 115(47.1) | 98(40.2) | 11.12 | 0.085 |
| Literate (informally) | 52 | 7(13.5) | 27(51.9) | 18(34.6) |  |  |
| Primary school | 25 | 6(24.0) | 8(32.0) | 11(44.0) |  |  |
| Secondary and above | 39 | 3(7.7) | 12(30.8) | 24(61.5) |  |  |
| **Wealth quintile, n(%)** |  |  |  |  |  |  |
| Lowest | 72 | 11(15.3) | 30(41.7) | 31(43.1) | 10.10 | 0.258 |
| Second | 72 | 8(11.1) | 34(47.2) | 30(41.7) |  |  |
| Middle | 72 | 11(15.3) | 27(37.5) | 34(47.2) |  |  |
| Fourth | 72 | 5(6.9) | 32(44.4) | 35(48.6) |  |  |
| Highest | 72 | 12(16.7) | 39(54.2) | 21(29.2) |  |  |
| **Financially dependent on children, n(%)** | 92 | 10(10.9) | 33(35.9) | 49(53.3) | 8.07 | 0.089 |
| No | 137 | 16(11.7) | 70(51.1) | 51(37.2) |  |  |
| Partially | 131 | 21(16.0) | 59(45.0) | 51(38.9) |  |  |
| Totally |  |  |  |  |  |  |
| **Main occupation, n(%)** | 258 | 31(12.0) | 120(46.5) | 107(41.5) | 10.95 | 0.027 |
| Agriculture | 51 | 3(5.9) | 26(51.0) | 22(43.1) |  |  |
| Employed/business | 51 | 13(25.5) | 16(31.4) | 22(43.1) |  |  |
| Housework |  |  |  |  |  |  |
| Abbreviations: SD, standard deviation; BMI, body mass index; MNA, mini nutritional assessment. |  |  |  |  |  |  |
| <sup>a</sup>ANOVA test for mean differences between nutritional status groups. <sup>b</sup>F test statistics. All others are Chi-square tests. |  |  |  |  |  |  |

Individuals with farming as their occupation were significantly more often malnourished (p=0.02). Wealth quintile and financial dependency were not significantly associated with malnutrition in the study.

### Health related characteristics and Nutritional status

Table 4 shows the results of analyses for health-related characteristics and nutritional status. Regarding health-related characteristics, individuals having insomnia and hospitalization during the last year were more often experiencing malnutrition. No significant association was demonstrated between independent variables such as having more chronic illnesses, multiple drug intake, and having chronic pain, and nutritional status.

**Table 4.** Nutritional status and associated health related characteristics.

| Variables | Total<br>(n=360) | Malnourished<br>47, 13.1% | Risk of<br>malnutrition<br>162, 45.0% | Well<br>nourished<br>151, 41.9% | Test<br>statistics | p-value<sup>c</sup> |
| --- | --- | --- | --- | --- | --- | --- |
| **Number of diseases,**<br>**n(%)** | 348 | 46(13.2) | 153(44.0) | 149(42.8) | 4.58 | 0.101 |
| <=2, | 12 | 1(8.3) | 9(75.0) | 2(16.7) |  |  |
| >2 |  |  |  |  |  |  |
| **Daily drug intake,**<br>**n(%)** | 316 | 39(12.3) | 139(44.0) | 138(43.7) | 3.43 | 0.179 |
| <=3, | 44 | 8(18.2) | 23(52.3) | 13(29.5) |  |  |
| >3 |  |  |  |  |  |  |
| **Chronic pain, n(%)** |  |  |  |  |  |  |
| Yes | 67 | 10(14.9) | 37(55.2) | 20(29.9) | 5.00 | 0.082 |
| No | 293 | 37(12.6) | 125(42.7) | 131(44.7) |  |  |
| **Insomnia, n(%)** |  |  |  |  |  |  |
| Always/Often | 103 | 29(28.2) | 51(49.5) | 23(22.3) | 39.08 | 0.000 |
| Occasionally/No | 257 | 18(7.0) | 111(43.2) | 128(49.8) |  |  |
| **Hospitalization during one year, n(%)** |  |  |  |  |  |  |
| Yes | 48 | 13(27.1) | 21(43.8) | 14(29.2) | 10.52 | 0.005 |
| No | 312 | 34(10.9) | 141(45.2) | 137(43.9) |  |  |
| **Oral health, n(%)** |  |  |  |  |  |  |
| **Chewing problem** |  |  |  |  |  |  |
| Yes | 167 | 26(15.6) | 78(46.7) | 63(37.7) | 3.03 | 0.220 |
| No | 193 | 21(10.9) | 84(43.5) | 88(45.6) |  |  |
| **Edentulous** |  |  |  |  |  |  |
| Yes | 301 | 40(13.3) | 141(46.8) | 120(9.9) | 3.35 | 0.187 |
| No | 59 | 7(11.9) | 21(35.6) | 31(52.5) |  |  |

<sup>c</sup>All statistics are Chi-square tests.

## Discussion

This study assessed the nutritional status of community-dwelling older individuals and socio-demographic factors, health status, and social indicators on nutrition. The study revealed a high prevalence of poor nutritional status and health status among older people. The main findings of the present study were 13.1% malnutrition and 45.0% at risk of malnutrition among the elderly, as measured by MNA. Illiteracy, having more co-morbidities, and having insomnia were all associated with poor nutritional status.

### Nutritional status

The present study revealed a prevalence of 13.1% malnutrition and 45.0% at risk of malnutrition among the elderly. BMI data showed 11.7% underweight elderly population, similar to MNA malnutrition. A study in a village of Nepal assessed malnutrition using MNA and showed 24% of elderly were malnourished and 65% were at risk of malnutrition (15). A cross sectional study in a rural VDC of Nepal revealed malnutrition 11.6% of population were malnourished and 49.7% were at risk of malnutrition (19). This findings of this study are similar to our study.

Sabita et al. (20) found 19.8% malnourished and 45.7% at risk of malnutrition among elderly using MNA tool in urban areas of terai region of Nepal. A study carried out in villages nearby Kathmandu valley, Pharping VDC, among elderly population using MNA revealed that 31% of elderly people were malnourished and 51% were at risk of malnutrition (36). The lower malnutrition rate seen in this study may be due to the urban population of elderly people in the study.

In a rural population of South India, a study reported a prevalence of malnutrition similar to this study. The study used MNA to assess the nutritional status among older persons aged 60 years and above and found a prevalence of malnutrition of 14% and at risk of malnutrition of 49%. The current study showed a similar prevalence of malnutrition. In a cross-sectional study among rural community-dwelling elderly Lebanese, Beulos et al. (10) found a prevalence of 8% malnutrition and 29.1% at risk of malnutrition. These results are close to the present study. The current study showed a lower rate of malnutrition compared with the findings from a study of rural older persons of Bangladesh (9). In this sample, MNA was used to assess the nutritional status, and the prevalence of malnutrition was 26%, and 62% were at risk of malnutrition.

The nutritional status assessed by MNA in developed countries showed a lower prevalence of malnutrition in community-dwelling older adults. In a review of 24 studies with more than 30,000 older adults worldwide, Guigoz et al (24) reported that the mean prevalence of malnutrition among community-dwelling elderly was 1%. A higher prevalence was found in outpatients (4%), hospitalized elderly (20%), and among elderly in institutional homes (37%). Similarly, Kaiser et al. (34) found a prevalence of malnutrition of 5.8% and that 31.9% were at risk of malnutrition in a pooled result of data from five countries, including 964 community-dwelling elderly individuals. In this analysis, the authors found 22.8% malnutrition and 46.2% at risk of malnutrition in 24 combined datasets (hospital, nursing home, community, and rehabilitation settings), including information on more than 6000 elderly study participants from different settings, including home-living older adults.

In developing countries, poor nutrition is a prevalent condition among the elderly. In Nepal, a cross-sectional study (33) revealed 13% malnutrition, measured by BMI (BMI<18.5), among elderly home-living older individuals. However, the use of BMI as the only tool to detect malnutrition becomes unreliable (32).

The high prevalence of malnutrition or risk of malnutrition indicates the multidimensionality of malnutrition in old age in Nepal (2). Research has shown that multiple complex factors are in interplay for the development of poor nutrition (30). The present study also supports the previous findings.

### Socioeconomic factors

Old age itself is a risk factor for malnutrition, as has been reported in the literature. Anorexia of aging is a cause of malnutrition in older persons, including other factors such as multiple pathologies, functional impairment, and decreased food intake (14, 37). Anorexia of aging causes decreased appetite and decreased feeding, leading to malnutrition (6). The present study found a significant association between increasing age and malnutrition. Our study showed almost doubled malnutrition (20%) in those aged 80 years and above compared to 10% among those aged 60 to 70 years. This is consistent with other previous studies.

The results also indicate that low educational status was associated with poor nutritional status. In the study, the illiteracy rate was 68%, and women were found to be two times more illiterate compared to men. Similar findings were reported in a previous study (10). Better education has been positively correlated with good nutritional status (31, 38).

Women had more malnutrition than men in previous studies. Low education, frequent widowhood, and increased comorbidity among women may explain this finding (35). However, the current study does not support the association of malnutrition and gender.

Being financially dependent on children is a risk factor for malnutrition, as demonstrated in previous studies. Higher income was positively correlated with nutritional status. Boulos et al (5) states “economic factors are drivers of nutritional status.” Another unexpected finding in the current study is that financial dependency did not reveal any significant association with malnutrition. A possible reason may be that more than 60% of the elderly live with children in Nepal (17).

Tamang et al. (27) found increased malnutrition approximately 25% in elderly in a study in eastern Terai region of Nepal. Unemployment, more daily use of prescribed medication and comorbidity were significantly associated with malnutrition in this study which is similar to our study findings. Notable findings in our study were the lack of association of gender, ethnicity, marital status, occupation, and oral health status with the nutritional status of the elderly.

### Health status

Nearly 65% of the study population had at least one chronic disease. Hypertension was the most prevalent disease (26.5%), followed by Diabetes Mellitus (12.5%), COPD/Asthma (12.0%), and arthritis. One study in Nepal found 59.3% of participants reported chronic pain, 35% reported respiratory problems, and 22% had hypertension (40). This finding is comparable to the current study. One-third of the participants reported their health status as good according to SRH. A cross-sectional study in Bangladesh (31) found that all of the participants were suffering from at least one medical condition (31). In a study, most of the elderly had some form of disease diagnosed by a doctor or health worker (39). Various studies have reported that increased comorbidity is associated with malnutrition (5, 9, 32). Increased comorbidity was associated with malnutrition in this study.

Chronic pain is also implicated in malnutrition, since 18% of the population had chronic pain. Pain causes loss of appetite and loss of pleasure, leading to poor nutrition (5). Interestingly, chronic pain was also not significantly associated with malnutrition in this study, although several studies have reported an association of poor nutritional status with chronic pain, chronic diseases, and an increased number of daily drug intakes (27, 31). About 12.2% of individuals reported taking more than 3 prescribed medicines daily.

Nearly 34% were currently smoking tobacco. Regarding physical exercise, 25% of the participants were regularly exercising, and there was no significant difference between men and women on smoking and physical exercise as seen in some studies (5, 27). There was no significant association of malnutrition with smoking and exercise in this study.

About 28.6% reported having insomnia always or often. Insomnia was significantly associated with malnutrition in this study. Approximately 28% of elderly individuals with insomnia were malnourished. Ferede et al. (29) found similar association of insomnia and malnutrition among elderly in their study. About 13.1% of the individuals were hospitalized within one year, and recent hospitalization had increased malnutrition in the elderly.

### Limitation of the study

Due to its cross-sectional nature, causality could not be determined. Various measures were self-reported status, so recall bias may have occurred in the study. The study population is in urban areas on the outskirts of Lalitpur metropolitan city of Nepal, and may not represent the population of the rural areas of Nepal.

## Conclusions

This is a study to describe the prevalence of nutritional status, health-related characteristics, and social indicators in an urban community-dwelling elderly population in Nepal. The present study provided unique information regarding the nutritional status, health status, and social indicators of community-dwelling elderly people in urban areas of Lalitpur, Nepal.

Malnutrition was found in 13% of elderly people, and 45% were at risk of malnutrition. Poor nutritional status was a common condition among old people, and this study described the proximal and distal determinants of poor nutritional status among the elderly in Nepal. Since elderly malnutrition is often undetected and underestimated, elderly nutritional screening should be incorporated in national health survey. This study indicates an urgent need to take care of the health of the elderly for successful aging with timely nutritional and health interventions.

## Recommendations

Further research on a national representative sample is required to identify the factors associated with poor nutritional status.

A longitudinal research is recommended to understand the direction of the causality of the findings. Involvement of multi-sector providers should be promoted to enhance nutritional status in old people. Health care providers should reinforce the knowledge of the elderly regarding nutrition and health.

## Acknowledgements

We would like to thank the Department of Public Health, Maharajgunj Medical Campus, Nepal, Prof. Dr. Sharad Raj Onta for guiding the research, and the participants, without whom this study would not have been possible.

## Author contributions

Conceptualization: Eebaraj Simkhada, Aerusha Simkhada

Formal analysis: Eebaraj Simkhada

Investigation: Eebaraj Simkhada, Aerusha Simkhada

Supervision: Eebaraj Simkhada.

Writing – original draft: Eebaraj Simkhada.

Writing – review & editing: Eebaraj Simkhada, Aerusha Simkhada

## Data Availability Statement

Data of the study are available on request from the corresponding author (Eebaraj Simkhada).

